# Clinical and molecular response of AML harboring non-canonical *FLT3* N676K driver mutations to contemporary FLT3 inhibitors

**DOI:** 10.1101/2022.09.15.22279953

**Authors:** Gregory W. Roloff, Frank Wen, Aubrianna Ramsland, Andrew S. Artz, Satyajit Kosuri, Wendy Stock, Olatoyosi Odenike, Richard A. Larson, Hongtao Liu, Lucy A. Godley, Michael J. Thirman, Anand A. Patel, Christopher K. Daugherty, Adam S. DuVall, Mariam T. Nawas, Emily Dworkin, Geoffrey D. Wool, Carrie Fitzpatrick, Jeremy P. Segal, Peng Wang, Michael W. Drazer

**Affiliations:** Section of Hematology/Oncology, The University of Chicago, Chicago, IL; Division of Hematology & Hematopoietic Cell Transplantation, City of Hope, Duarte CA; Department of Pathology, The University of Chicago, Chicago IL

## Abstract

The treatment of acute myeloid leukemia (AML) has been enhanced by the development and regulatory approval of a series of novel agents, including midostaurin and gilteritinib (FLT3 inhibitors), venetoclax (BCL2 inhibitor), ivosidenib (IDH1 inhibitor), and enasidenib (IDH2 inhibitor). A difficulty that has arisen in the era of molecular therapies, however, is determining the efficacy of these agents for patients with AML harboring atypical driver mutations. The non-canonical FLT3 p.N676K variant was initially described as an acquired resistance mechanism in patients with FLT3 internal tandem duplication (ITD) mutations who were treated with midostaurin. Clinical data from patients with the FLT3 N676K mutations are limited. Here, we detail our experience caring for nine different patients with AML harboring FLT3 N676K at the University of Chicago. Seven of nine (78%) individuals received intensive induction chemotherapy, with FLT3 inhibitors utilized in three patients upfront and five patients during subsequent lines of treatment. With the use of FLT3 inhibitors, we noted reduction, and in some instances, complete molecular suppression of detectable FLT3 N676K variant allele fraction (VAF) on NGS, underscoring the activity of FLT3 inhibitors in this population regardless of line of therapy. Individuals with FLT3 N676K-mutated AML who received FLT3 inhibitors had longer median survival (940 days) than those who did not (408 days), however, the difference was not significant likely due to small size of the study (p = 0.2). The presence of concurrent canonical FLT3 mutations was associated with loss of treatment response. In silico visualization models of the FLT3 tyrosine kinase domain in the presence of gilteritinib demonstrate that the mechanism of N676K-mediated resistance is not due to disruption of FLT3 inhibitor binding at the ATP-binding site but rather influenced by other allosteric forces on protein structure. In conclusion, this is the largest study to date demonstrating that the atypical FLT3 N676K driver mutation is sensitive to contemporary FLT3 inhibitors, such as midostaurin and gilteritinib. Our data suggest that FLT3 inhibitors should be included in both the upfront induction setting as well as in the relapsed/refractory setting for patients harboring the FLT3 N676K mutation.

---

Treatment of acute myeloid leukemia (AML) has been enhanced by the development and regulatory approval of several novel agents, including midostaurin and gilteritinib (FLT3 inhibitors), venetoclax (BCL2 inhibitor), ivosidenib (IDH1 inhibitor), and enasidenib (IDH2 inhibitor) (1). A difficulty during the era of molecular therapies, however, is determining the efficacy of these agents for patients with AML harboring atypical driver mutations. These atypical drivers were underrepresented in seminal clinical trials that led to the approval of targeted AML therapies, thereby limiting availability of data for clinical decision making (2). The non-canonical *FLT3* N676K variant was initially described as an acquired resistance mechanism in patients with *FLT3* internal tandem duplication (ITD) mutations treated with midostaurin (3). *In vivo* studies demonstrated *FLT3* N676K-mutated AML is sensitive to midostaurin and quizartinib, but suggested that cooperating ITD mutations confer resistance to both agents (4). Clinical reports of *FLT3* N676K-mutated AML are limited to those of two individuals, both of whom developed *FLT3* N676K mutations at relapse (4, 5). Treatment outcomes for *de novo* disease with *FLT3* N676K mutations are lacking, and limited data exist regarding the utility of FLT3 inhibitors for *FLT3* N676K-mutated AML patients.

The aim of this study was to use clinical and genomic data to investigate the efficacy of FLT3 inhibitors for *FLT3* N676K-mutated AML. We performed a retrospective analysis of patients with AML receiving care at the University of Chicago. The study was approved by the Institutional Review Board and conducted according to the Declaration of Helsinki. Our practice utilizes a validated 1213 gene next-generation sequencing (NGS) panel that has been previously described (6-8). NGS is employed at presentation and at subsequent time points to assess response or disease status. In cases of morphologic remission, NGS is not performed due to the anticipated lack of detectable tumor DNA.

We identified nine patients with AML and *FLT3* N676K mutations. N676K was the only *FLT3* mutation detected in seven patients, whereas two patients had co-incident ITD or tyrosine kinase domain (TKD) mutations at some point during their clinical course. The median age at AML diagnosis was 41 years (17-79 years), with a mean presenting WBC count of 53 300/μL. Four of nine patients had normal cytogenetics. Laboratory and clinical data can be found in Table 1.

**Table 1.** Clinical and laboratory characteristics of patients with *FLT3* N676-mutated AML.

|  | Pt 1 | Pt 2 | Pt 3 | Pt 4 | Pt 5 | Pt 6 | Pt 7 | Pt 8 | Pt 9 |
| --- | --- | --- | --- | --- | --- | --- | --- | --- | --- |
| Sex | F | F | M | M | F | F | F | M | M |
| Age Range | 46-50 | 66-70 | 40-45 | 65-70 | 50-55 | 36-40 | 36-40 | 25-30 | 15-20 |
| Presenting WBC (k/uL) | 70 | 1.5 | 87.8 | 123 | 93 | 25 | 15.4 | 2.8 | 60.2 |
| Presenting BM Blasts (%) | 56 | 37 | 65 | n/a | n/a | 74 | 95 | 80 | 78 |
| Presenting PB Blasts (%) | 5 | 0 | 43 | 92 | 76 | 68 | 90 | 0 | 80 |
| Cytogenetics | t(8;16) | 46, XX | 46, XX | n/a | +8 | 46, XX | n/a | del(13q) | 46, XY |
| *FLT3* mutation at diagnosis | N676K | None | None | N676K | ITD N676K | D835V N676K | n/a | n/a | N676K |
| Induction backbone | 7+3 | Enasidenib | 7+3 | None | 7+3 | 7+3 | 7+3 | 7+3 | 7+3+ Etoposide |
| Frontline FLT3i | Sorafenib | None | None | None | Midostaurin | Midostaurin | None | None | None |
| Induction response | CR MFC MRD- | PD | CR | None | CR *NPM1* MRD- | CR *NPM1* MRD- | CR | CR MFC MRD- | PD |
| *FLT3* mutation at relapse | none | N676K | N676K | None | ITD | N676K | N676K | N676K | N676K |
| Subsequent FLT3i? | Midostaurin | Gilteritinib | Midostaurin | None | Gilteritinib | None | None | None | Gilteritinib |
| Setting for subsequent FLT3i | Maintenance | Salvage & Post-HCT maintenance | Post-HCT maintenance | None | Maintenance | None | None | None | Salvage |
| Best Response | CR MFC MRD- | CR MFC MRD- | CR *NPM1* MRD- | n/a | CR *NPM1* MRD- | CR *NPM1* MRD- | CR MFC MRD- | CR MFC MRD+ | CR MFC MRD- |
| HCT? | Yes | Yes | Yes | No | No | Yes | No | Yes | Yes |
| Alive? | Yes | Yes | Yes | No | No | No | No | No | No |
| Survival (days)* | NR (217) | NR (1407) | NR (2864) | 4 | 445 | 516 | 239 | 577 | 1363 |

Abbreviations used: Pt, patient; F, female; M, male; WBC, white blood count; BM, bone marrow; PB, peripheral blood; FLT3i, FLT3 inhibitor; HCT, hematopoietic cell transplant; n/a, not available/not applicable; PD, progressive disease; NR, not reached; MFC, multiparameter flow cytometry; MRD, measurable residual disease. \*For living patients, survival calculated as of manuscript submission and included in parentheses.

To infer antileukemic activity of FLT3 inhibitors for *FLT3* N676K-mutated AML and to characterize relapse dynamics, we analyzed *FLT3* N676K variant allele frequency (VAF) kinetics in patients for whom longitudinal NGS data were available. Regardless of disease status at the time of FLT3 inhibitor use (*de novo* vs relapse), patients receiving FLT3 inhibitors had declines in *FLT3* N676K VAF. For individuals in whom *FLT3* N676K was the only *FLT3* mutation (Patients 1, 2, 9), a mean treatment time of 95 days led to undetectable *FLT3* N676K. Suppression of *FLT3* N676K VAF generally paralleled clinical response and likelihood of survival at the time of our analysis (Fig. 1A). One exception was patient 9, who died of hepatic veno-occlusive disease after gemtuzumab ozogamicin treatment and subsequent allogeneic transplantation despite having morphologic and molecular remission with gilteritinib-based salvage therapy.

**Figure 1.**
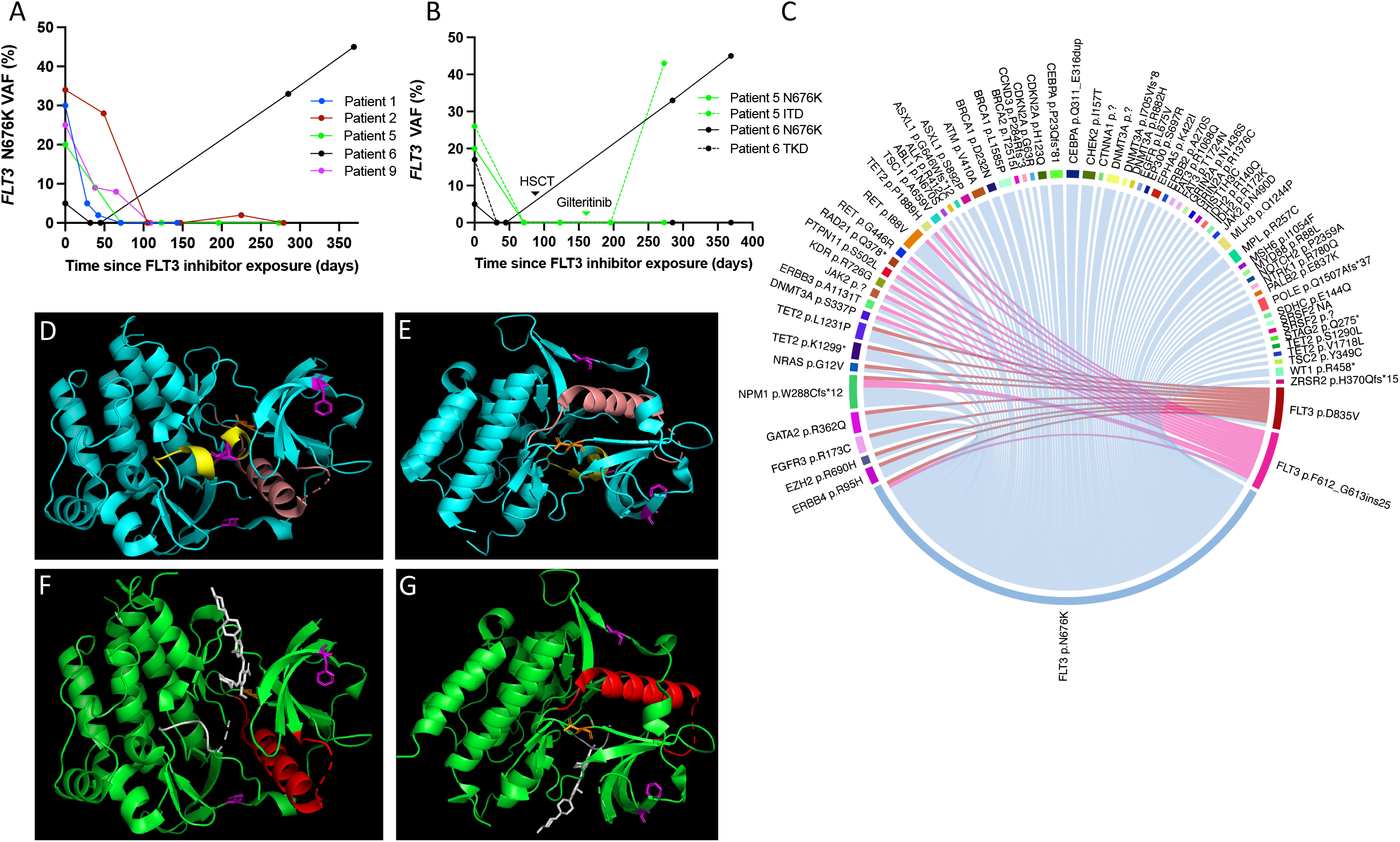
**(A)** Longitudinal detection of *FLT3* N676K by next-generation sequencing from the time of FLT3 inhibitor initiation. **(B)** Clinical course and relapse dynamics of two patients with *FLT3* N676K-mutated AML and coincident ITD or TKD mutations. (**C**) Circos plot of comutational networks of *FLT3* N676K-mutated AML patients in the analysis. Chord thickness reflects the number of co-occurrences between two genes. (**D)** PyMol visualization models of the uninhibited FLT3 tyrosine kinase domain with N676K (residue shown in orange) and activation loop (yellow). Residues F612 and D835 (corresponding to ITD and TKD mutations, respectively) are shown in magenta. (**E**) Upon activation, the FDG-containing alpha helix (salmon) translocates inward. Inhibition by gilteritinib at the ATP-binding site is depicted in the inactive, FDG-out (**F**) and activated, FDG-in (**G**) state with the FDG-containing alpha helix shown in red.

One older patient pursued comfort care immediately after diagnosis. Six patients were treated with “7+3” induction therapy, with 3/6 patients having FLT3 inhibitors added to induction chemotherapy (midostaurin, n=2; sorafenib, n=1). Patient 1 had a complete morphologic and molecular remission. They proceeded to allogeneic transplantation after induction 7+3 therapy with sorafenib (discontinued for gastrointestinal toxicity) and subsequent midostaurin during pre-transplant consolidation. Day 30 post-transplant bone marrow studies demonstrated a complete morphologic and molecular remission on midostaurin maintenance therapy.

Two additional patients (patients 5 and 6) harbored *de novo* disease with co-occurring *FLT3* mutations, one with a *FLT3* ITD mutation (patient 5) and the other a *FLT3* TKD mutation (patient 6). Each had midostaurin added to 7+3 induction. By days +71 and +32 after midostaurin treatment, respectively, both demonstrated remission with no detectable *FLT3* VAF (Fig. 1B).

Patient 5 had *de novo* disease with co-occurring *FLT3* mutations (*FLT3* ITD F612_G613ins25 and N676K). She received 7+3+midostaurin induction and had a morphologic and molecular complete remission 71 days after initiating midostaurin. Induction was complicated by fungal pneumonia and repeated episodes of acute kidney injury. She was not a candidate for cytotoxic consolidation therapy and started gilteritinib. She continued to experience multiple episodes of acute kidney injury unrelated to gilteritinib. Gilteritinib was held during these episodes, and she eventually presented with 25% circulating blasts after approximately 100 days of intermittent gilteritinib administration. NGS at relapse showed ascendancy of the same *FLT3* ITD clone (VAF 43%) that was present at diagnosis, but an absence of the *FLT3* N676K mutation. The patient chose comfort measures.

Patient 6 had *de novo* disease with co-occurring *FLT3* TKD (D835V) and N676K mutations. She underwent induction with 7+3+midostaurin, which led to morphologic and molecular remission 32 days after initiating midostaurin. She proceeded to hematopoietic cell transplantation in first remission. Despite MRD negativity at transplant, she relapsed after 6 months. She did not receive post-transplant FLT3 inhibitor therapy. NGS performed at relapse demonstrated expansion of the *FLT3* N676K population (VAF 33%) and absence of the original *FLT3* TKD clone. Salvage measures with donor lymphocyte infusion and high dose cytarabine/mitoxantrone were unsuccessful. She died of complications from central nervous system leukemic infiltration.

We also analyzed the spectrum of other pathogenic mutations coexisting with *FLT3* N676K in our cohort (Supplementary Fig. 1). Co-mutational clusters were most notable for *FLT3* N676K and either *FLT3* ITD or *FLT3* TKD mutations (Fig. 1C). To understand the structural properties of therapeutic inhibition of FLT3 in the presence of the N676K mutation, we utilized PyMOL (Schrödinger) to study the FLT3 TKD harboring N676K in the presence and absence gilteritinib (9). Upon activation, three residues, Asp-Phe-Gly (DFG), shift inward (DFG-in) from the inactive state (DFG-out). Mutations at D835 within the TKD favor the active DFG-in state and promote resistance to type II FLT3 inhibitors (10). Gilteritinib and other type I FLT3 inhibitors bind directly to the ATP-binding site, maintaining their activity regardless of DFG conformation (11). As shown in Fig. 1 D-G, the N676K mutation does not prohibit the transition from DFG-out to DFG-in or the interaction of gilteritinib with the ATP-binding site.

A recent analysis of the mutational landscape of patients with *FLT3*-mutated AML treated on CALGB 10603 (RATIFY) showed 26/275 (5.5%) of patients harbored non-canonical *FLT3* mutations (2). Ten of these 26 patients (38%) had *FLT3* N676K-mutated AML (2). Growing clinical application of NGS will increase the identification of atypical driver mutations. Robust clinical series focused on *FLT3* N676K-mutated AML patients are lacking, and the benefit of FLT3 inhibitor therapies in this population was previously unknown.

Here, we used clinical and genomic data to assess the utility of FLT3 inhibitors in the largest series of *FLT3* N676K AML patients described to date. Although previously described to be enriched in populations of core binding factor AML (4), none of the seven patients in our cohort with metaphase cytogenetic data available had core-binding factor AML. Seven of nine (78%) individuals received intensive induction chemotherapy. FLT3 inhibitors were utilized in three patients during frontline induction and five patients during subsequent lines of therapy. We observed reduction, and in some instances, complete molecular suppression of detectable *FLT3* N676K VAF on NGS, underscoring the activity of FLT3 inhibitors in this population, regardless of the line of therapy. All patients with *FLT3* N676K mutations who were treated with FLT3 inhibitors had a best response of MRD negativity via flow cytometry or NGS at some point during their care.

Consistent with previous evidence (3, 4), concurrent canonical ITD or TKD *FLT3* mutations were associated with loss of treatment response. *In silico* modeling of FLT3 in the presence of gilteritinib suggests that the mechanism of N676K-mediated resistance is not due to disruption of FLT3 inhibitor binding at the ATP-binding site but is likely influenced by other allosteric forces on protein structure. Individuals with *FLT3* N676K-mutated AML in our cohort whose treatment included FLT3 inhibitors had longer median survival (940 days) than those who did not (408 days, excluding patient 4 who immediately pursued comfort measures). This difference was not significant, likely because of the small size of this study (*p* = 0.2). The three patients who remained in an ongoing remission at the time of manuscript submission, however, were all treated with FLT3 inhibitors. With emerging evidence supporting the role of post-transplant FLT3 inhibitor maintenance therapy for suppression of *FLT3* ITD-mutated AML (12), further study evaluating the durability of FLT3 inhibitor maintenance for patients with non-canonical driver mutations in both transplant and non-transplant settings is warranted.

In conclusion, this is the largest study to date demonstrating that the atypical *FLT3* N676K driver mutation is sensitive to contemporary FLT3 inhibitors, such as midostaurin and gilteritinib. This mutation has been infrequently detected in seminal studies of FLT3 inhibitors. However, our data demonstrate FLT3 inhibitors should be included both in upfront induction setting and relapsed/refractory settings for patients harboring the *FLT3* N676K mutation.

## Data Availability

All data produced in the present study are available upon reasonable request to the authors

## AUTHOR CONTRIBUTIONS

MWD designed the study; MWD and GWR collected the data; MWD, GWR, FW, and AR analyzed the data; MWD, GWR, ASA, SK, WS, OO, RAL, HL, LAG, MJT, AAP, CKD, ASD, MTN, and ED cared for the patients described. GDW, CF, JPS and PW were responsible for pathologic analysis and molecular database curation. All authors edited the manuscript and agreed with the final version.

