## Supplementary Materials for "Clinical and molecular response of AML harboring non-canonical *FLT3* N676K driver mutations to contemporary FLT3 inhibitors"

Table of Contents:

**Figure S1.** Spectrum of Pathogenic NGS variants across patient cohort **Figure S2.** Comutational correlation analysis of longitudinal NGS data

**Figure S3.** Intra-patient mutation profiles detected on longitudinal NGS.

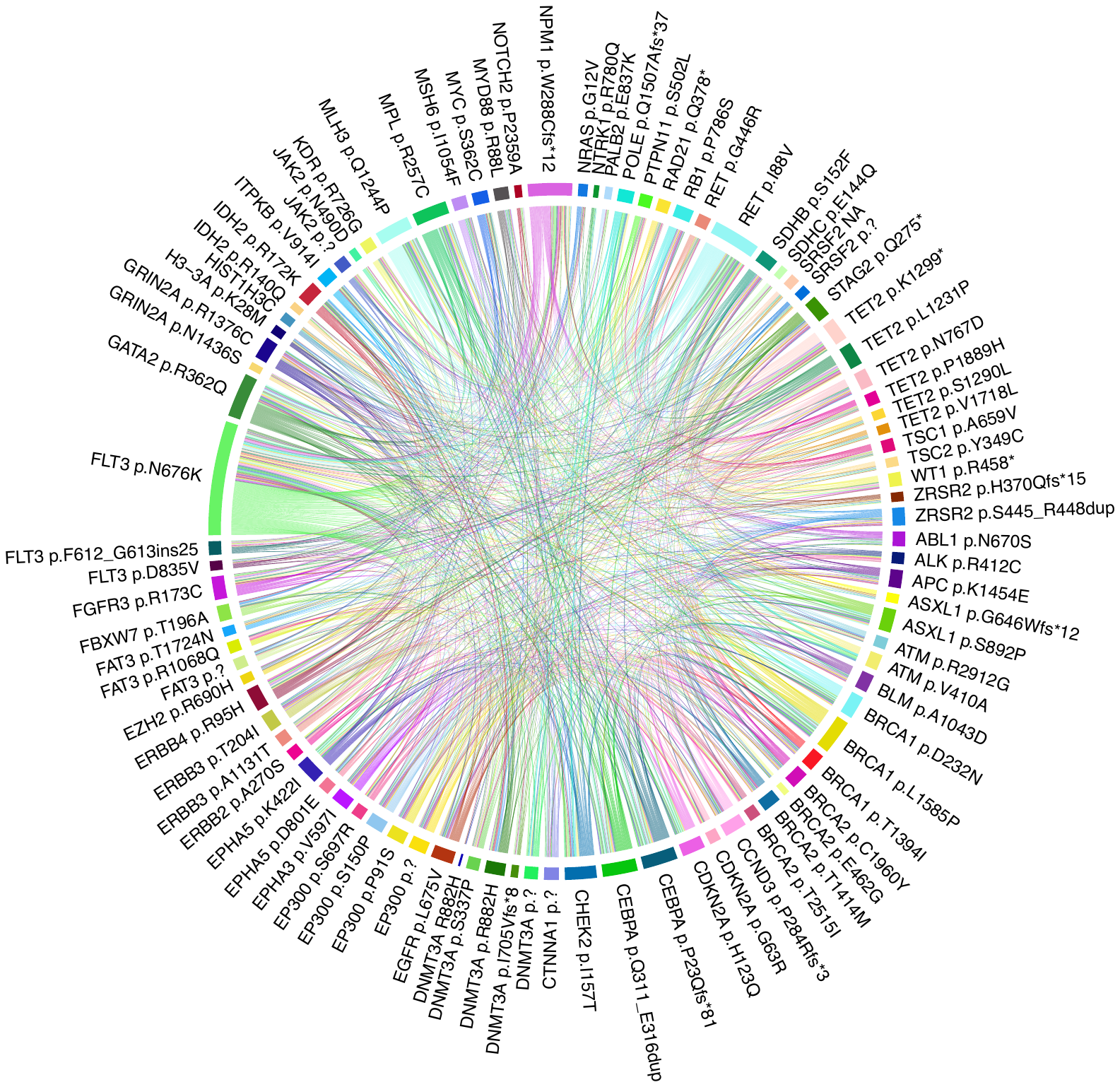

**Figure S1. Spectrum of Pathogenic NGS variants across patient cohort.**

Circos plot showing the complete spectrum of NGS-detected variants across the FLT3 N676K patient cohort. A total of 185 individual mutations were observed. Width of the circle diameter for associated genes (shown in lime green for FLT3 N676K) corresponds to mutational frequency.

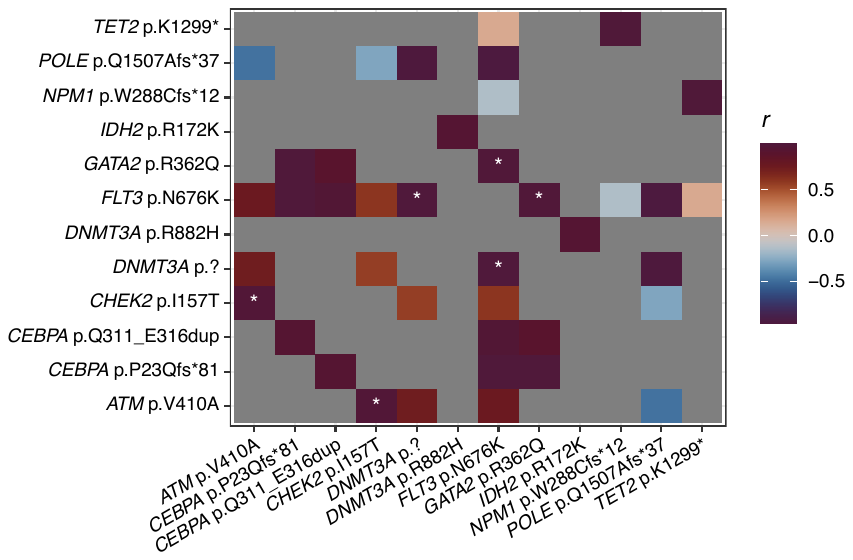

**Figure S2. Comutational correlation analysis of longitudinal NGS data.**

Pearson’s correlation coefficient was across variant allele frequencies (VAFs) for all observed instances of comutations that co-occur at least twice in the dataset. Values approaching 1.0 on the heatmap indicate a strong propensity for mutational co-occurance while more negative values denote mutual exclusivity between two mutational events. White asterisks indicate p < 0.05.

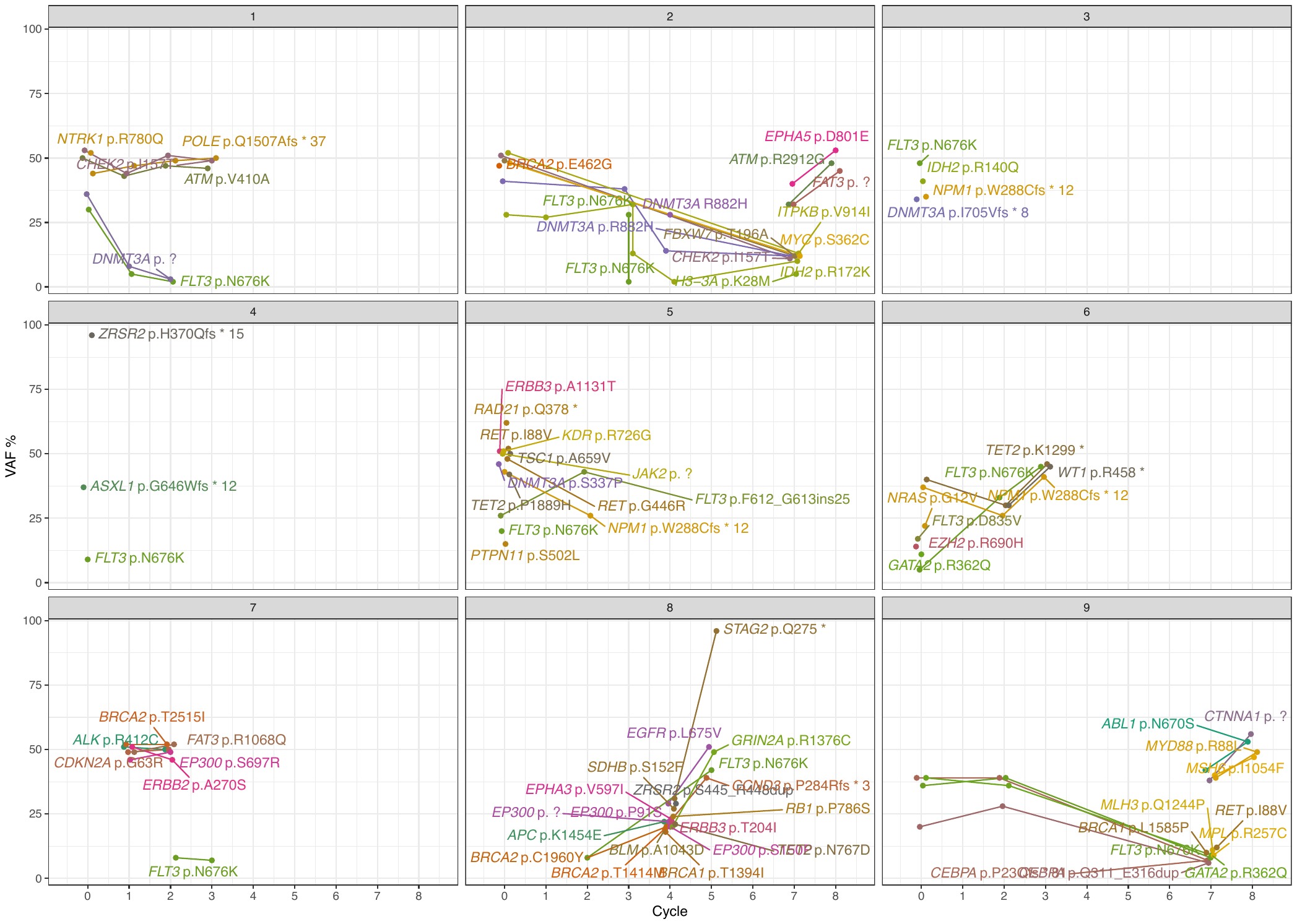

**S3. Intra-patient mutation profiles detected on longitudinal NGS.**

Longitudinal NGS demonstrates VAF kinetics over time. Each cycle indicated on the x-axis represents a repeat NGS assessment over the clinical course.
